# Baseline Mismatch Negativity Amplitude Predicts Direction and Magnitude of Ketamine Effect in Healthy Volunteers — A “Disordinal” Effect

**DOI:** 10.1101/2025.10.07.25337443

**Authors:** M. Cecchi, J. Johannesen, B. Farley, M.C. Quirk, M. Mahmoud-Zadeh, J.M. Uslaner, R. Terry-Lorenzo, D.G. Smith, D.A. Ruhl, M. Rotte, A.L. Reese, P. O’Donnell, J. Mollon, C. Missling, Y. Matsuoka, M.J. Marino, S. Lee, I.O. Korolev, D. Klamer, A. Jeong, S. Honda, K.C. Fadem, J. Doherty, E.A. Cohen, S. Christensen, H. Chadchankar, D.L. Buhl, M. Adachi, D.C. D’Souza, H. K. Hamilton, M. Ranganathan, B.J. Roach, L. Ereshefsky, D.P. Walling, W.Z. Potter, D.C. Javitt, D.H. Mathalon

## Abstract

**Background:** Mismatch negativity (MMN) is a component of the auditory event-related potential (ERP) that is elicited during a passive oddball paradigm where task-irrelevant infrequent deviants are presented in a stream of more frequent standard stimuli. MMN is believed to index a pre-attentive stage of auditory information processing closely linked to N-methyl-D-aspartate receptors (NMDAR). Ketamine is thought to act primarily as an NMDAR antagonist, has been used in clinical trials to model the symptoms of schizophrenia and is increasingly used in the clinic to treat depression. Various studies have reported that ketamine reduces MMN amplitude which, in turn, might reflect reduced function of NMDAR-mediated neurotransmission. Nonetheless, there is growing evidence showing MMN amplitude either having high variability or, paradoxically, moving in the opposite direction after ketamine in different individuals.

**Methods:** In here, we analyzed results from three independent ERP studies to test the hypothesis of a cross-over interaction (“disordinal” drug effect) between the duration-deviant MMN at baseline (without ketamine) and the direction and magnitude of the ketamine effect. To rule out regression to the mean (RTM), a statistical phenomenon that may also partially explain this cross-over interaction, we separately estimated RTM using a drug-free test-retest study.

**Results:** Our results are the first to statistically demonstrate the existence of a disordinal drug response to ketamine, where the direction and magnitude of ketamine-induced changes in MMN amplitude can be predicted by baseline MMN amplitude.

**Conclusions:** These new insights may contribute to novel precision medicine approaches to treatment of CNS disorders.

## Introduction

Recent efforts in the pharmaceutical field to advance precision medicine for neuropsychiatric conditions have focused on identifying biomarkers that can index drug effects on brain function. One promising modality is electroencephalography (EEG) and EEG-based event-related potentials (ERP).

Mismatch negativity (MMN) is an EEG response elicited during a passive auditory oddball ERP paradigm, where an occasional “deviant” sound is presented in a stream of more frequent “standard” sounds. MMN is a negative deflection in the difference wave generated by subtracting the ERP to the standard stimulus from the ERP to the deviant stimulus (Figure 1). MMN indexes a pre-attentive stage of auditory information processing and has been linked to NMDAR function as well as excitation/inhibition (E/I) balance involving interactions between glutamatergic (excitatory) pyramidal and GABAergic (inhibitory) local circuit neurons (1,2).

**Figure 1.**
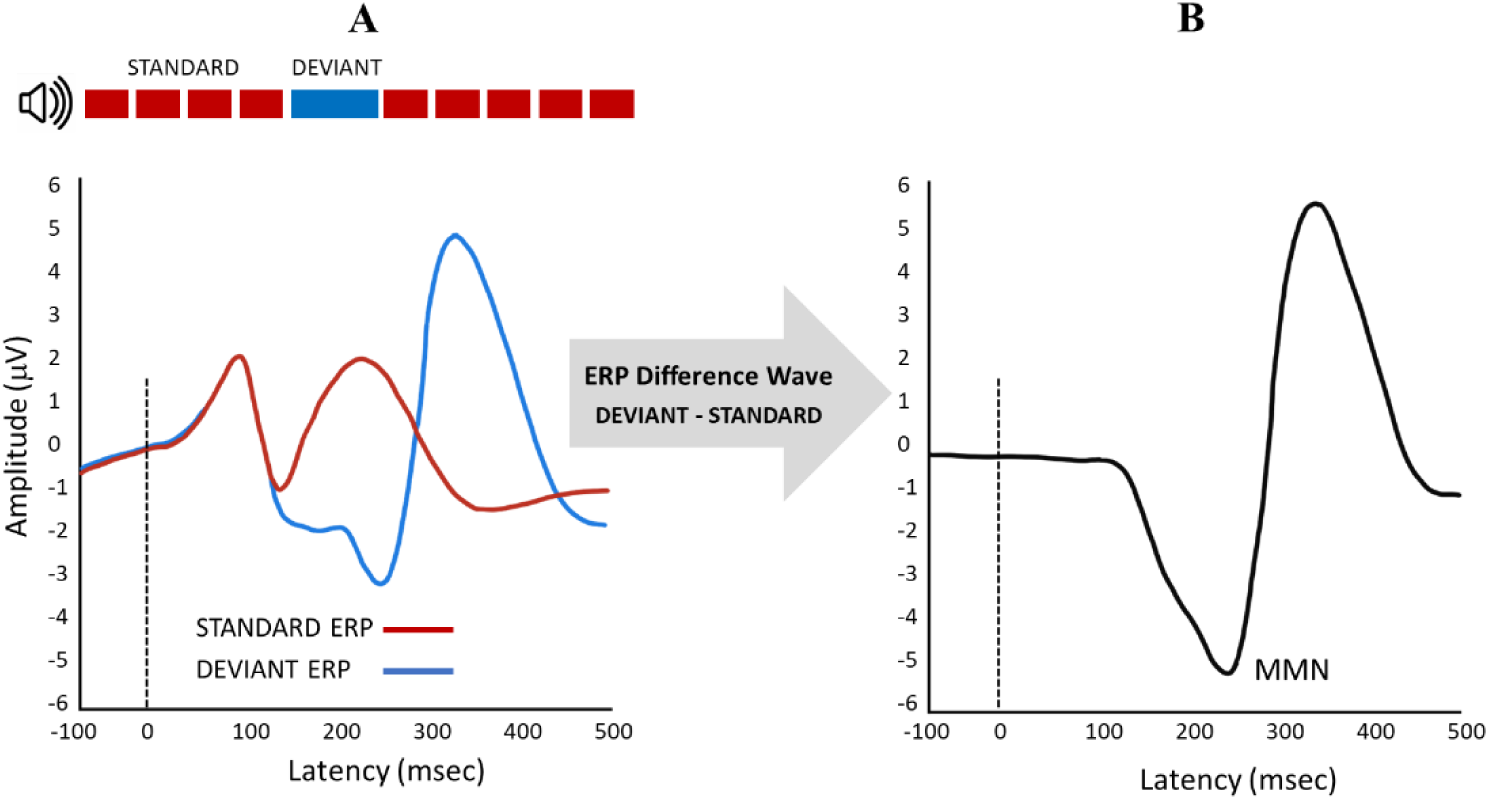
MMN from a Passive Auditory Oddball ERP Paradigm. (A) shows a MMN paradigm and the ERP to Standard sounds (red) and Deviant sounds (blue). (B) shows the resultant ERP difference wave after the Standard ERP is subtracted from the Deviant ERP. The MMN component is labeled in the ERP difference wave as the negative wave peaking around 230 msec post-stimulus onset.

Though numerous studies have reported that NMDAR antagonists such as ketamine reduce MMN amplitude (MMNa) (3,4), recent work suggests that the link between NMDAR function and MMNa is not as clear (see for example (5)). In fact, during a review of pharmaceutical company sponsored clinical trials where we collected EEG during a passive auditory oddball paradigm under no-ketamine (NK) and ketamine-on-board (KOB) conditions, we noted that ketamine did not attenuate MMNa in all study participants. Our studies (NCT03771586, NCT03844906, NCT05049343, NCT04928703) included a total of 128 healthy subjects across 280 total NK and KOB testing sessions, which to our knowledge represents the largest series of ketamine-challenge studies using substantially equivalent methods. Our results indicated a bidirectional effect on MMNa where the direction and magnitude of the response to ketamine was correlated with the NK MMNa. We have termed this cross-over interaction a “disordinal effect” of ketamine on MMNa (6).

In this paper, we analyze pooled results from three independent MMN studies to statistically confirm our observation that ketamine has a disordinal effect on MMNa. A key factor in this analysis is a required discrimination between ketamine’s possible disordinal effect and an effect due to “regression-to-the-mean” (RTM). RTM is evident when the change in test scores from one session to another (i.e., change score) correlates with the single session test scores from which change was calculated. Typically, the single session is the first session (i.e., “baseline”) and RTM reflects the change in the distribution of subjects from the first to the second session that correlate with the first session’s distribution, reflecting the familiar “baseline correlation with change” effect (although RTM can also be evident retrospectively, with the distribution of scores in the second session correlating with the change scores). In this RTM-driven relationship, the subjects with more extreme test scores at the first session have scores closer to the mean at the second session.

We illustrate these relationships with hypothetical MMN data collected over two sessions (Test 1, Test 2) in Figure 2A, with a regression line showing a negative correlation between Test 2 – Test 1 MMN change scores and Test 1 MMN scores and no overall change in the means from Test 1 to Test 2. This negative correlation indicates that subjects with more negative (i.e., larger) MMN scores at Test 1 tend to have less negative (i.e., smaller) MMN scores at Test 2 (i.e., positive change scores), and subjects with less negative (i.e., smaller) MMN scores at Test 1 tend to have more negative (i.e., larger) MMN scores at Test 2 (i.e., positive change scores). In Figure 2B, we illustrate a hypothetical scenario where ketamine administered at Test 2 shows a uniform unidirectional effect of reducing MMN scores (i.e., less negative mean MMN) that superimposes on the RTM effect. Here, the ketamine-induced change relative to placebo (i.e., Test 2 – Test 1) is reflected by a positive shift in the intercept of the regression line (“positive mean effect”), with no change in the slope of the regression line relative to the slope of the RTM regression line in Figure 2A. In Figure 2C, we illustrate the hypothetical scenario where ketamine administered at Test 2 has a true disordinal effect over and above the RTM effect. In this case, regression of the ketamine induced MMN change (Test 2 – Test 1) on placebo (Test 1) MMN scores results in a negative correlation with a significantly steeper slope than that resulting from RTM alone. As shown in Figure 2C, this steeper slope reflects the disordinal effect such that subjects with larger (i.e., more negative) MMN scores on placebo tend to show a greater reduction in MMN (i.e., MMN is less negative) with ketamine (positive effect), whereas subjects with smaller (i.e., less negative) MMN scores on placebo tend to show a greater increase in MMN (i.e., MMN is more negative) with ketamine (inverse effect). This hypothetical disordinal effect is similar to what has been previously referred to as Wilder’s Law of Initial Value, which states that the magnitude and direction of the effect of a stimulus on a physiological or psychological measure are influenced by the baseline level of the measure prior to the stimulus (7).

**Figure 2.**
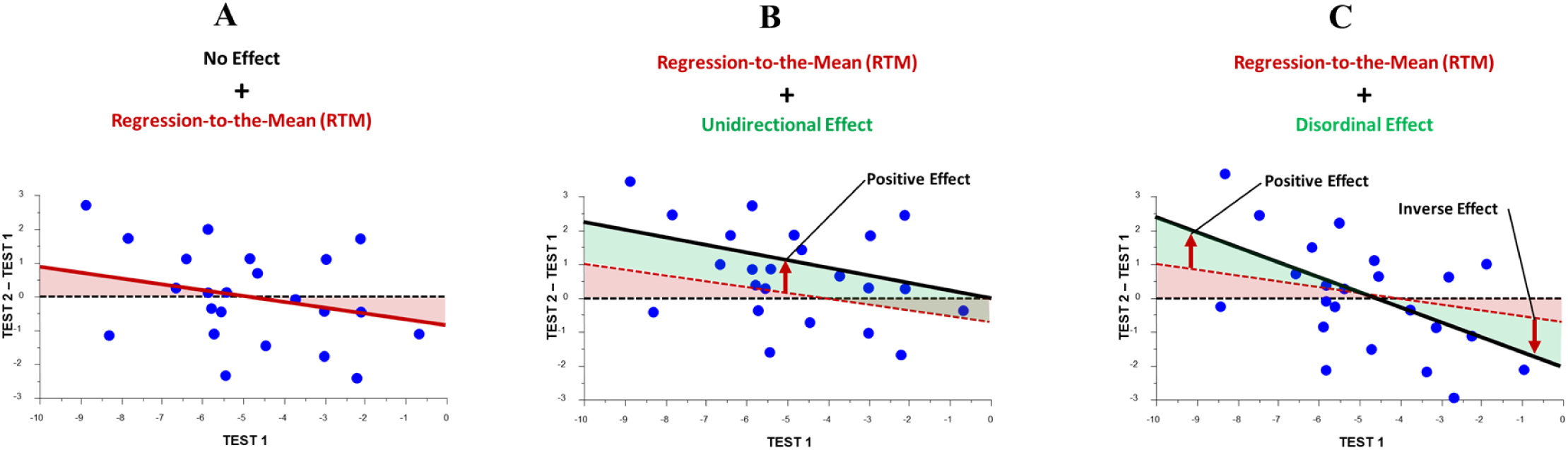
Hypothetical regression line slopes illustrating regression to the mean (RTM) vs unidirectional ketamine effect vs disordinal ketamine effects on MMN amplitude. Examples of test-retest data showing linear regression lines for MMN change scores (Test 2 – Test 1) regressed on MMN baseline (Test 1) scores under different hypothetical scenarios. (A) shows a RTM effect when the TEST1 and TEST2 are the same subjects tested twice under identical conditions (no drug). The RTM effect is evident as a negative slope of the regression line (red solid line) relating MMN change scores (Test 2 – Test 1) to MMN Test 1 scores. (B) shows a unidirectional effect of ketamine (Test 2) relative to placebo (Test 1) superimposed on the RTM effect. The ketamine-induced MMN change score (Test 2 – Test 1) correlation with baseline (i.e., placebo) TEST1 scores has the same slope as the one induced by RTM alone (red dotted line), indicating RTM with no additional disordinal effect due to ketamine. Here, ketamine exerts a unidirectional “positive mean effect” on MMN baseline scores (i.e., MMN becomes smaller, or less negative, with ketamine), reflected by a positive shift in the intercept of the regression line relative to the RTM regression line as indicated by the green shading. (C) shows a disordinal effect of ketamine (Test 2) relative to placebo (Test 1) over and above the RTM effect (red dotted line). This disordinal effect is reflected by a negative correlation between ketamine-induced change in MMN (Test 2 – Test 1) and baseline placebo MMN (Test 1) that has a significantly steeper slope (black solid line) than the one associated with RTM alone (red dotted line). This steeper slope reflects a greater reduction in MMN (i.e., less negative MMN) by ketamine in those subjects with larger (i.e., more negative) baseline MMN scores (i.e., “positive effect” of ketamine), and a greater increase in MMN (i.e., more negative MMN) by ketamine in those subjects with smaller (i.e., less negative) baseline MMN scores (i.e., “inverse effect” of ketamine). The disordinal effect, shown in green shading at both ends of the Test 1 score distribution, is evident over and above the RTM effect shown in red shading.

In the current study, we estimate the correlation between baseline placebo MMN scores and ketamine-induced change scores from three different ketamine vs. placebo challenge studies to determine whether a consistent inverse relationship is present. Then, to determine whether this inverse relationship is a true disordinal effect of ketamine, the slope of this relationship is compared to the slope of the simple RTM-driven baseline vs. change score relationship evident from a separate study of healthy volunteers tested twice over a 1 week interval with no drug administered (NCT04025502)(8). We further examine the mean change in MMN due to time alone, likely a reduction in MMN due to habituation to the task over repeated sessions, relative to the mean change in MMN produced by ketamine across the 3 ketamine challenge studies included in the current report, using meta-analytic methods.

## Methods and Materials

### Ketamine-Challenge Studies

We analyzed results from three double-blind, placebo-controlled clinical trials in healthy volunteers who underwent at least one MMN session under NK conditions and one MMN session under KOB conditions.

The first study (KET1, n=24, NCT04928703) was a 3-arm, 3-period crossover design. Study participants were administered IV ketamine in two of the sessions and placebo in the remaining session in a counterbalanced randomized order. For KET1, comparisons of treatment arms showed no significant session order effects on MMN amplitudes, including a lack of difference between the two MMN_KOB_ sessions irrespective of session order. Accordingly, the average MMN amplitude across the two active ketamine sessions representing MMN_KOB_, along with the MMN amplitude from the placebo session representing MMN_NK_, were included in our analyses.

The second study (KET2, n=19, NCT05049343) was a Phase 1, crossover trial of SAGE-904 as a ketamine block. This study dosed participants with SAGE-904 or placebo, followed by ketamine administration. Only data from the placebo (MMN_NK_) and placebo-ketamine (MMN_KOB_) sessions, completed in the morning and afternoon of the same day, respectively, were included in our analyses.

The third study (KET3, n=27) was a counterbalanced crossover design that examined the interactive effects of ketamine and nicotine to determine whether nicotine prevents or attenuates MMN abnormalities (9). In this study, no effects of session order were evident on the MMN amplitudes for placebo and active ketamine days. Only the placebo and ketamine sessions were included in our analyses, irrespective of session order. While this study demonstrated significant reductions of MMN by ketamine for pitch, intensity, and pitch+duration double deviants, ketamine did not significantly reduce duration-deviant MMN.

#### Ketamine Administration

For all studies, ketamine was administered intravenously as a 0.23mg/kg bolus over 1 minute, followed by an initial maintenance infusion at 0.58mg/kg/hour for 30 minutes, and a reduced infusion rate of 0.29mg/kg/hour after that for up to 64 minutes. This ketamine dose is in the same range as what is commonly recommended for treatment of depression (10), and the dosing protocol has been shown to achieve stable ketamine plasma levels for the duration of ketamine administration (11).

### Regression-to-the-Mean Study

The study used to quantify the RTM slope for MMNa in healthy volunteers (RTM1, n=35, NCT04025502) was sponsored by the ERP Biomarker Qualification Consortium (www.erpbiomarkers.org). This 4-site observational study included healthy volunteers and participants with schizophrenia who were tested across two sessions (Time1 & Time2) one week apart. No drug was administered at either session. This study was specifically designed to quantify test-retest variances for EEG/ERP measures, including MMNa (8). Only healthy volunteer data were included in our analyses.

### Study Participants

All studies were approved by institutional review boards, and written informed consent was obtained from each study participant. Demographics for the participants included in the analyses are shown in Table 1.

**Table 1.** Demographics for study participants included in the analyses.

|  | *RTM1* | *KET1* | *KET2* | *KET3* |
| --- | --- | --- | --- | --- |
| Sample Size | 35 | 24 | 19 | 27 |
| Age<sup>1</sup> | 38.11 (1.91) | 33.75 (1.07) | 42.56 (2.06) | 25.78 (1.20) |
| Sex |  |  |  |  |
| Male<sup>2</sup> | 17 (48.6%) | 19 (79.2%) | 13 (68.4%) | 15 (55.6%) |
| Female<sup>2</sup> | 18 (51.4%) | 5 (20.8%) | 6 (31.6%) | 12 (44.4%) |
| Race |  |  |  |  |
| White<sup>2,3</sup> | 8 (22.9%) | 3 (12.5%) | 8 (42.1%) | 21 (77.8%) |
| African American<sup>2,3</sup> | 4 (11.4%) | 19 (79.2%) | 11 (57.9%) | 1 (3.7%) |
| Other Race<sup>2,3</sup> | 23 (65.7%) | 2 (8.3%) | 0 (0.0%) | 5 (18.5%) |
| Education<sup>1</sup> | 13.83 (.32) | 12 (0.00) | n/a | 15.52 (0.51) |
| BMI<sup>1</sup> | n/a | 25.87 (0.62) | 27.21 (0.50) | 23.88 (0.96) |

Abbreviations: BMI= body mass index. n/a = not available.
Notes:
1. Mean ( ± SEM).
2. Total (% of Total).
3. Racial labels from FDA, 2016, Guidance for Industry, Collection of Race and Ethnicity Data in Clinical Trials (12).

### Mismatch Negativity Data Acquisition and Processing

#### MMN Paradigm

For KET1, KET2, and RTM1, a duration-deviant MMN testing paradigm was administered as described in Cecchi et al., 2023 (8). For KET3, the duration-deviant MMN was elicited as part of a multi-deviant paradigm similar to Näätänen et al., 2004 (9,13).

In all cases, duration-deviant stimuli were 50ms longer than standards. All study participants were instructed to ignore auditory stimuli and perform a picture-name verification task (14) to minimize the influence of attention on MMN.

#### Electroencephalography Data Acquisition, Preprocessing, and Extraction of MMN Amplitude

For KET1, KET2, and RTM1, electroencephalography (EEG) data were recorded during the MMN paradigm using a commercially available COGNISION^®^ System (Cognision, Louisville, KY, USA). Data cleaning, preprocessing and feature extraction were automatically performed using a processing pipeline implemented in COGNISION^®^ Software, as previously described in Cecchi et al, 2023 (8).

For KET 3, EEG data were recorded during the MMN paradigm using Neuroscan Synamps amplifiers and an EasyCap electrode cap (Brain Products GmbH, Gilching, Germany). For additional details about data acquisition, preprocessing, and feature extraction, see Hamilton et al, 2018 (9).

#### Z-Score Normalization

Separately for each study, MMN amplitudes for each session were z-score normalized with respect to the mean and standard deviation (SD) of the study’s NK session (for KET studies) or Time1 session (for RTM1) before statistical analyses were conducted. Specifically, within each study, and for MMN data from each session, the mean MMN_NK_ or MMN_RTM1_ was subtracted from each participant’s raw MMN amplitude, and this deviation score was then divided by the MMN_NK_ or MMN_RTM1_ SD to yield a z-score. As a result, within each study, MMN_NK_ or MMN_RTM1_ z-scores had mean=0 and SD=1. In effect, all MMN amplitudes within each study were re-expressed as z-scores reflecting deviations, in standard units, from the mean values observed during NK or RTM1 test session. In this way, variation between the studies, including in hardware, recording environments, processing pipelines, or healthy participant samples, were minimized. All reported MMNa analyses are performed using these MMN z-scores.

### Analytical Design and Statistical Analysis

To assess the degree to which “baseline” placebo MMN amplitude scores (i.e., MMN_NK_) are correlated with the change in MMN amplitude produced by ketamine (i.e., MMN_KOB_ – MMN_NK_), MMN change scores were regressed on MMN_NK_ scores, Study (KET1, KET2, KET3), and the Study x MMN_NK_ interaction. Here the focus was on testing the interaction effect to determine whether the three studies showed inverse relationships with similar slopes, reflecting a common potential disordinal effect of ketamine.

Next, to test whether the expected inverse relationship between ketamine induced change in MMN and baseline MMN was a true disordinal relationship, the three ketamine studies were combined to derive a single estimate of the inverse relationship that was then statistically compared with the RTM inverse relationship. This was done in a second regression model in which MMN change scores were regressed on baseline MMM scores, Study (Ketamine vs RTM), and the Study x Baseline MMN interaction. Here, the test of the interaction, reflecting whether regression line slopes significantly differed between ketamine and RTM studies, was the critical test of whether a true disordinal effect of ketamine was evident over and above the RTM effect. We also provide tests of the change vs. baseline Pearson correlations within each study to address whether each study’s regression line slope significantly differed from zero.

In addition to testing disordinal effects vs. RTM1 effects using regression models, we compared ketamine’s effect sizes on mean MMN amplitude z-scores across each of the three ketamine challenge studies with each other and with the effect size of the MMN change due to time alone using a meta-analytic random-effects model and estimates of the 95% confidence intervals for each study’s effect size. Separate subgroup analyses were performed for KET1, KET2, KET3 and RTM1 studies to estimate effect sizes for each study. Cohen’s *d*, representing the standardized mean difference, was used as the measure of effect size. A forest plot was generated to visualize individual study effect sizes, pooled effect size, and heterogeneity statistics.

Further, for differences in slopes between the RTM1 study and the combined KET studies, we conducted the Johnson-Neyman procedure (15) to identify the range of MMNa z-score values where the change induced by ketamine is significantly larger or significantly smaller than the change in MMN attributable to RTM alone.

For all comparisons, threshold for statistical significance was set at *p* = 0.05.

## Results

### Slope Comparisons among Ketamine Studies

Regression of ketamine-induced MMN z change scores (KOB-NK) on baseline (NK) MMN z-scores, Study (KET1, KET2, KET3), and their interaction revealed a trend for the regression line slopes to differ between studies (*F*_2,64_= 3.081, *p* = 0.053). Follow-up univariate Pearson correlations indicated that MMNa z change-scores were significantly associated with MMN_NK_ for KET1 (*b* = -0.500, 95% CI [-0.81, -0.19], *p* = 0.003; *r* = 0.580) (Figure 3-A), KET2 (*b* = -1.244, 95% CI [-1.78, -0.71], *p* < 0.001; r = 0.764) (Figure 3-B), and KET3 (*b* = -0.910, 95% CI [-1.34, -0.48], *p* < 0.001; *r* = 0.659) (Figure 3-C). Further pairwise comparisons between studies showed that the negative slope for KET1 showed a trend toward being smaller than for KET2 (*t*_64_= 1.898, *p* = 0.062), with no other slope differences between the ketamine studies. Dropping the interaction term from the model yielded a significant common slope (*b* = - 0.859, *p* < 0.001) across all three ketamine studies, which is similar to the mean of the three slopes (*b* = -0.883). Based on these results, the MMN z-score data from the three ketamine studies were combined to yield one large ketamine vs. placebo sample for comparison with the data from the RTM1 study.

**Figure 3.**
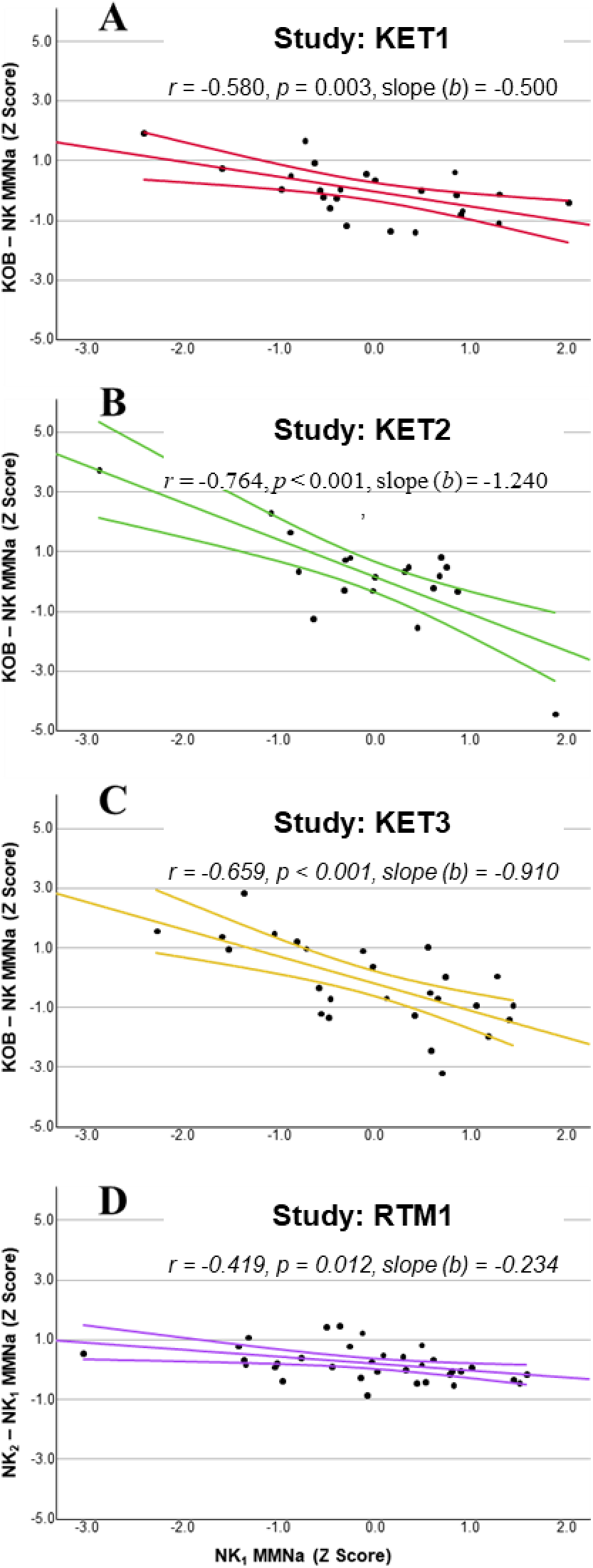
Ketamine Effect Linear Regressions for 3 KET Studies and Regression to the Mean (RTM1) Study. Linear regressions for MMN z-scores for KOB - NK for the ketamine (KET) studies as well as for NK2 – NK1 for the RTM1 study versus baseline MMN z-scores. Regression lines are linear fits with confidence intervals (95% of the mean).

### Slope Comparisons Between Ketamine and RTM1 Studies

In the regression of MMN z change scores (KOB-NK for combined ketamine studies, NK2 – NK1 for RTM1 study) on baseline MMN z-scores, Study (KET vs. RTM1), and their interaction revealed a significant interaction effect (*F*_1,101_ = 11,824, *p* < 0.001) indicating significant slope differences between the studies. The RTM1 study showed a significant negative slope (*b* = -0.234, 95% CI [-0.41, -0.05], *p* = 0.012; *r* = -0.419), as shown in Figure 3D, indicating MMN z change-scores for RTM1 were significantly associated with Time1 MMN z-scores, consistent with a regression to the mean effect (Figure 3-D). Importantly, the slope in the combined ketamine study sample (*b* = -0.859, CI [-1.10, -0.62], *p* < 0.001, *r* = -0.653) was significantly more negative, consistent with a disordinal effect of ketamine on MNN over and above the RTM effect (see Figure 4).

**Figure 4.**
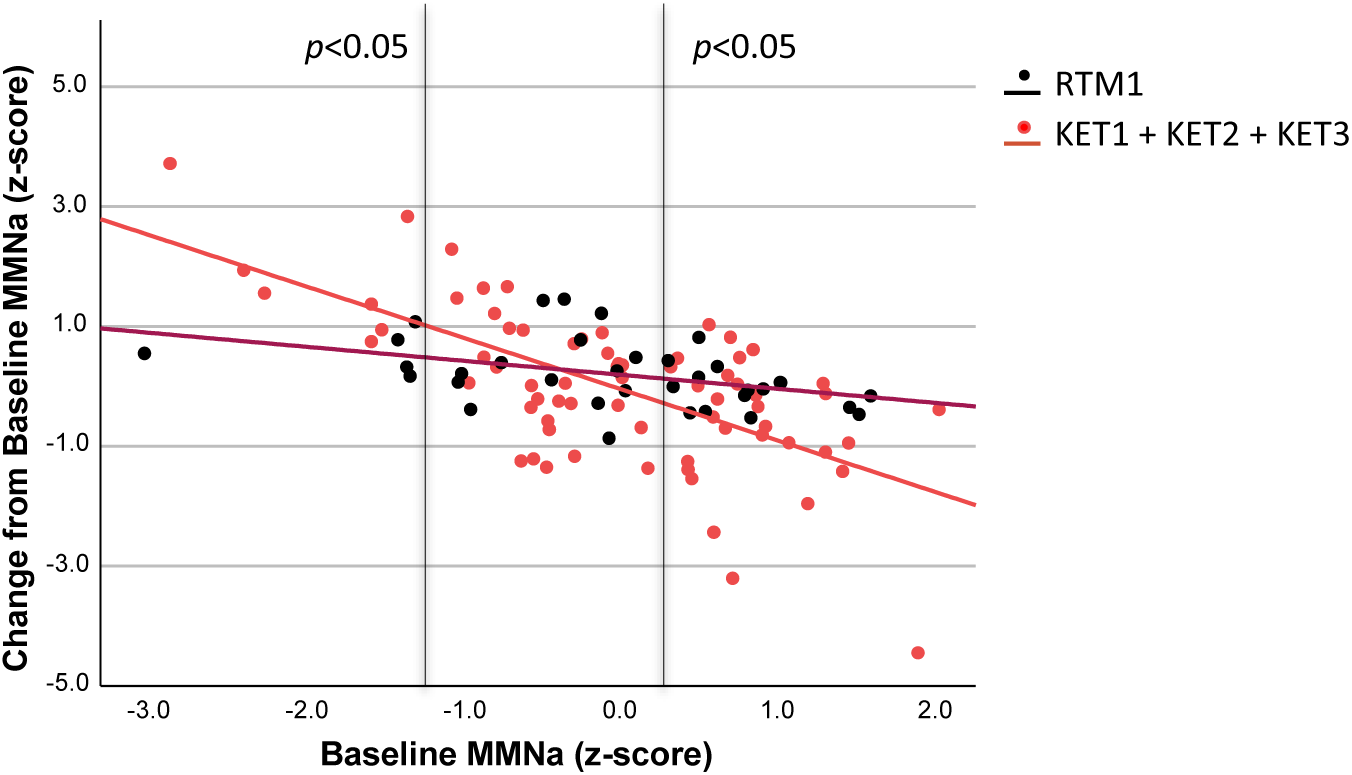
Ketamine Effect Linear Regression for the Combined ketamine (KET) Studies vs the Regression to the mean (RTM1) Study. Linear regression for MMN z-score change from baseline vs. baseline MMN z-scores for combined data from the 3 KET studies vs from the RTM1 study. Vertical lines identify the range of MMN z-score values where the change induced by ketamine was significantly different (p < 0.05) than the change due to regression to the mean per the Johnson-Neyman procedure.

A subsequent Johnson-Neyman procedure identified the range of baseline MMN z-scores for which the change induced by ketamine differed significantly from the change resulting from regression to the mean in RTM1. The change induced by ketamine was significantly larger (i.e., more negative) than regression to the mean for MMN baseline z-scores above 0.222 (43.81% of the sample), and significantly smaller (i.e., more positive) for baseline z-scores below -1.299 (10.48% of the sample).

### Across Study Comparisons

#### Ketamine Group Effects

The pooled effect size for all studies was small and not statistically significant (*d* = 0.049, *p* = 0.724, 95% CI [-0.222, 0.320]).

Separate subgroup analyses for the KET and RTM1 studies showed no significant effects in either group: d = -0.029, p = 0.864, CI [-0.361, 0.303] for the pooled KET studies, and d = -0.205, p = 0.393, CI [-0.265, 0.675] for RTM1 (Figure 5).

**Figure 5.**
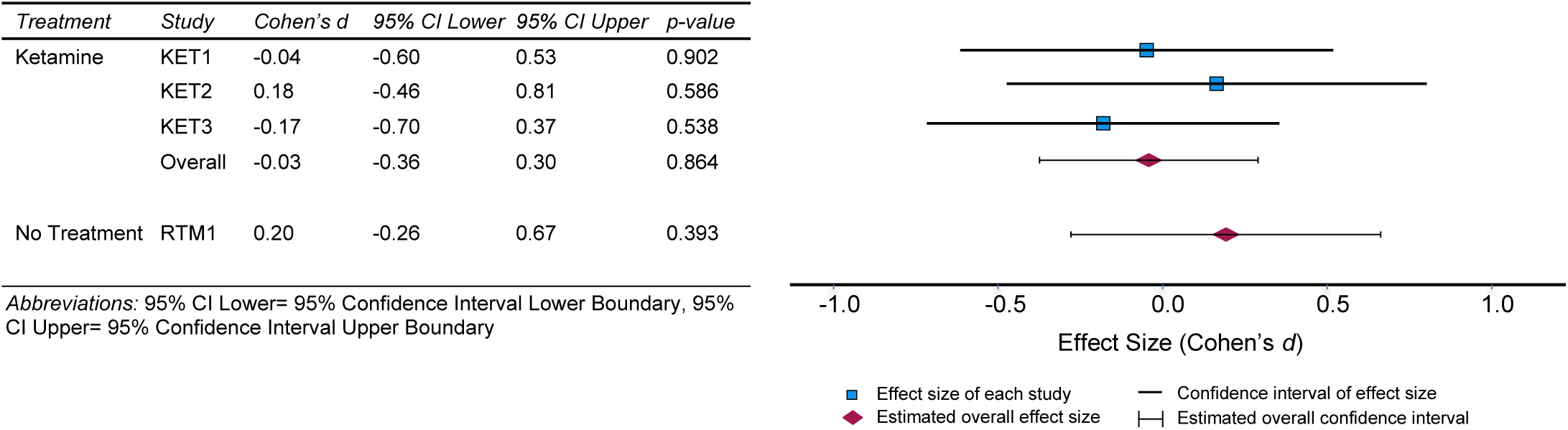
Ketamine Effect Sizes. Forest plot with effect sizes (Cohen’s *d*) and corresponding confidence intervals. Single study and pooled ketamine effect sizes are shown. A positive effect size indicates a smaller MMNa after ketamine for KET studies, and a smaller MMNa at Time 2 for RTM1.

Finally, heterogeneity across subgroups was low, as measured by the Cochran’s *Q* test (*Q* = 0.64, *p* = 0.43).

## Discussion

Our analyses show that, at the dose used in our studies, ketamine produces a “disordinal” effect distinct from RTM where the magnitude and direction of ketamine-induced changes on MMNa are dependent on the MMNa baseline value. Specifically, participants with large (i.e. more negative) baseline MMNa were most likely to experience ketamine-induced MMN reduction (“positive effect”; see Figure 2C), while participants with small (i.e., less negative) baseline MMNa typically experienced ketamine-induced MMN enhancement (“inverse effect”; see Figure 2C). As these data were collected across several different sites and with different hardware, these results may generalize for similar studies using MMNa as a measure of glutamatergic activity modulated by NMDAR-targeting compounds.

### Evidence of Disordinal Effects on MMNa

While to our knowledge this is the first report that statistically distinguishes a disordinal effect of a pharmacological intervention on MMNa from the baseline correlation with change effect associated with RTM, several published studies have reported results that are consistent with our findings. In a review on the role of glutamate transmission in MMN, Harms and coauthors report how in some cases NMDAR antagonism can increase MMNa, and that the effect is possibly modulated through inhibitory interneuron activity (16). In another recent review about the effects of neuroplastogens on brain function, the authors report that ketamine can increase the negativity of the MMN response to deviant tones, possibly indicating a heightened sensitivity to prediction error (17). Finally, in a study looking at the effects of memantine on MMNa in patients with chronic psychosis, Swerdlow and colleagues reported that the MMN-enhancing effects of memantine were most robust among patients with smaller MMNa during placebo administration (18).

A disordinal effect on MMNa may not be limited to compounds that directly affect the NMDAR. In a study that measured the acute effects of the α7 nAChR agonist cytidine 5′-diphosphocholine (CDP-choline) on MMNa in healthy volunteers, Knott et al. have reported a significant increase in MMNa in the low baseline amplitude group and the opposite effect in the high baseline amplitude group (19). The same lab reported similar results in patients with schizophrenia, suggesting that the disordinal effect may be maintained in clinical populations (20). One caveat here is that these studies did not demonstrate a disordinal effect on MMNa over and above the effect of RTM.

### Possible Mechanism for a Disordinal Response to Ketamine

The neurophysiological mechanisms underlying the ketamine-induced disordinal effect we observed on MMNa are unclear but likely to involve the intricate balance of excitatory and inhibitory signaling in cortical circuits mediated by the dynamic interplay between GABAergic and glutamatergic neurotransmission. Indeed, Javitt et al. have theorized that MMN represents selective current flow through the channels of cortical NMDA receptors that appear to be under tonic inhibition by GABAergic interneurons (1,2).

In addition, while a study investigating the effects of memantine on MMN in patients with frontotemporal lobar degeneration and age- and gender-matched controls found no significant overall effect on MMNa in either group, memantine-induced changes in MMNa correlated with patients’ prefrontal GABA concentration as measured by magnetic resonance spectroscopy (5). Specifically, in patients with low GABA concentration, MMNa was larger at baseline and memantine decreased the peak amplitude, whereas in patients with high GABA concentration, memantine moved MMNa in the opposite direction.

Recent studies also suggest that ketamine’s effect may be more complex than NMDA antagonism. Ketamine antidepressant effects appear to depend on the synergy between NMDA and opioid system activation (21) and could involve GABA and AMPA receptors (22). Thus, it is possible that ketamine’s effect on multiple neurotransmitter and receptor systems, including glutamatergic (via NMDA and AMPA receptors), GABAergic, and opioid systems, may mediate its disordinal effect on MMNa.

### Disordinal Effects on Other ERP Biomarkers

Disordinal effects of neuroactive drugs on ERP measures may not be limited to MMN responses. A study that looked at the effects of the D2 antagonist sulpiride on ERP measures from an active auditory oddball paradigm found that, although sulpiride had no significant effect on the amplitudes of the primary ERP components in the subject sample as a whole, it increased the P3b amplitude in low amplitude subjects and decreased it in high amplitude subjects (7).

Similarly, in a separate study using an active oddball paradigm, nicotine increased N100 and P3b amplitudes in participants with low baseline amplitude and had the opposite effect in subjects with high baseline amplitude (23).

While these studies described a possible disordinal effect, they did not demonstrate that the effect was greater than would be expected from RTM alone.

### Disordinal Effect vs. RTM

A commonly overlooked and/or poorly appreciated phenomenon in clinical trials is the effect of RTM on cross-over study designs or designs which include baseline vs change from baseline measures. The issue becomes most pronounced for study participants whose baseline scores fall in the tails of the distribution, as their follow-up scores will likely regress towards the mean in the absence of any intervention. If a study is adequately powered and uses an interventional agent that has a unidirectional effect on the biomarker, then RTM can usually be ignored. But as with ketamine, if the agent has a disordinal effect on the target measure, then a disordinal-vs-RTM analysis should be performed to distinguish the true drug effect from the effect of RTM alone.

### Implications of Disordinal Drug Effects in Precision Psychiatry Trials

A disordinal response to ketamine could account for weak or inconsistent effects of ketamine on MMN across studies, particularly for the duration deviant MMN that tends to show the weakest effect (3). The disordinal effect may also contribute to paradoxical patterns of results in late phase interventional trials of ketamine, esketamine, or similar compounds. Those trials have often failed to produce statistically significant effects (see for example NCT05414422). It is possible that a disordinal pattern of results on the study population could have obscured clinically important effects on a subset of patients.

Several groups have now begun programs to model the underlying mechanisms responsible for differences in MMNa response (see for example (24)). Armed with an understanding of disordinal ketamine effects, these efforts may ultimately lead to predictive algorithms to identify responders to these and similar treatments.

### Study Limitations

All studies included in the disordinal analysis in this manuscript used the same IV ketamine dose. Though in the same range as what is recommended for therapeutic intervention (10) and therefore relevant to drug development for clinical indications, it was the only dose tested. Testing of additional ketamine doses could inform on the dose-dependency of the disordinal effect.

All three studies considered in the analyses on the disordinal effect used a duration-deviant MMN. While we assume that a disordinal effect would be evident for other types of MMN, this will need to be examined in future studies.

Because data for other ERP parameters from the passive oddball paradigm are not available for one or more of the studies included in this report, our analyses were limited to the MMNa. Regression analyses for the P3a and N100 amplitude from the MMN paradigm for the KET1 study show slopes consistent with a simple RTM (data not shown), but additional studies will be required to definitively answer whether the observed disordinal effect in response to ketamine is specific to MMNa.

Finally, demographic factors could also affect the results and might be focus of further investigation, as well as determining if the disordinal effect would generalize across various clinical populations, such as schizophrenia and depression.

## Conclusions

Ketamine produces a “disordinal” effect on MMNa in healthy volunteers, inducing decreases in study participants with larger MMNa at baseline and increases in participants with smaller MMNa at baseline. The effect is distinct from regression-to-the-mean. These findings suggest the need for fundamentally new approaches to analyzing results of CNS interventional trials and for precision medicine in CNS indications.

## Data Availability

All data produced in the present study are available upon reasonable request to the authors

## Acknowledgments

Two of the studies included in the analyses (NCT04025502 and NCT04928703) were funded by the ERP Biomarker Qualification Consortium. A third study (NCT05049343) was funded by Sage Therapeutics. The fourth study was supported by AstraZeneca, the U.S. Department of Veterans Affairs and the Yale Center for Clinical Investigation.

This manuscript has been posted on a preprint server.

## References

1. Javitt D, Steinschneider M, Schroeder CE, Arezzo JC (1996): Role of cortical N-methyl-D-aspartate receptors in auditory sensory memory and mismatch negativity generation: implications for schizophrenia. Proc Natl Acad Sci U S A 93: 11962–11967.

2. Javitt DC (2024): Mismatch Negativity (MMN) as a Pharmacodynamic/Response Biomarker for NMDA Receptor and Excitatory/Inhibitory Imbalance-Targeted Treatments in Schizophrenia. Adv Neurobiol 40: 411–451.

3. Rosburg T, Kreitschmann-Andermahr I (2016): The effects of ketamine on the mismatch negativity (MMN) in humans - A meta-analysis. Clin Neurophysiol 127: 1387–94.

4. Schwertner A, Zortea M, Torres F V., Caumo W (2018): Effects of subanesthetic ketamine administration on visual and auditory event-related potentials (ERP) in humans: A systematic review. Front Behav Neurosci 12. 10.3389/fnbeh.2018.00070

5. Perry A, Hughes LE, Adams N, Naessens M, Murley AG, Rouse MA, et al. (2022): The neurophysiological effect of NMDA-R antagonism of frontotemporal lobar degeneration is conditional on individual GABA concentration. Transl Psychiatry 12. 10.1038/s41398-022-02114-6

6. Fadem K, Johannesen J, Farley B, Cecchi M, Ereshefsky L, Mathalon D (2023): Baseline Mismatch Negativity Amplitude Predicts Direction and Magnitude of Ketamine Effect in Healthy Volunteers: a “Disordinal” Effect. CNS Summitt, 2023 2.

7. Takeshita S, Ogura C (1994): Effect of the dopamine D2 antagonist sulpiride on event-related potentials and its relation to the law of initial value. International Journal of Psychophysiology 16: 99–106.

8. Cecchi M, Adachi M, Basile A, Buhl DL, Chadchankar H, Christensen S, et al. (2023): Validation of a suite of ERP and QEEG biomarkers in a pre-competitive, industry-led study in subjects with schizophrenia and healthy volunteers. Schizophr Res 254: 178–189.

9. Hamilton HK, D’Souza DC, Ford JM, Roach BJ, Kort NS, Ahn K-H, et al. (2018): Interactive effects of an N -methyl- d - aspartate receptor antagonist and a nicotinic acetylcholine receptor agonist on mismatch negativity: Implications for schizophrenia. Schizophr Res 191: 87–94.

10. Swainson J, McGirr A, Blier P, Brietzke E, Richard-Devantoy S, Ravindran N, et al. (2021, February 1): The Canadian Network for Mood and Anxiety Treatments (CANMAT) Task Force Recommendations for the Use of Racemic Ketamine in Adults with Major Depressive Disorder. Canadian Journal of Psychiatry, vol. 66. SAGE Publications Inc., pp 113–125.

11. Driesen NR, McCarthy G, Bhagwagar Z, Bloch MH, Calhoun VD, D’Souza DC, et al. (2013): The impact of NMDA receptor blockade on human working memory-related prefrontal function and connectivity. Neuropsychopharmacology 38: 2613–2622.

12. FDA (2016): Collection of Race and Ethnicity Data in Clinical Trials Guidance for Industry and Food and Drug Administration Staff Clinical Medical Preface Public Comment. Retrieved from http://www.regulations.gov.

13. Näätänen R, Pakarinen S, Rinne T, Takegata R (2004): The mismatch negativity (MMN): Towards the optimal paradigm. Clinical Neurophysiology 115: 140–144.

14. Perez VB, Ford JM, Roach BJ, Woods SW, McGlashan TH, Srihari VH, et al. (2012): Error monitoring dysfunction across the illness course of schizophrenia. J Abnorm Psychol 121: 372–387.

15. Hayes AF (2013): From Guilford Introduction to Mediation, Moderation, and Conditional Process Analysis AF2E. 7006: 9–10.

16. Harms L, Parras GG, Michie PT, Malmierca MS (2021): The Role of Glutamate Neurotransmission in Mismatch Negativity (MMN), A Measure of Auditory Synaptic Plasticity and Change-detection. Neuroscience 456: 106–113.

17. Agnorelli C, Spriggs M, Godfrey K, Sawicka G, Bohl B, Douglass H, et al. (2025): Neuroplasticity and Psychedelics: a comprehensive examination of classic and non-classic compounds in pre and clinical models. Neurosci Biobehav Rev 172: 106132.

18. Swerdlow NR, Bhakta S, Chou H-H, Talledo JA, Balvaneda B, Light G (2015): Memantine Effects On Sensorimotor Gating and Mismatch Negativity in Patients with Chronic Psychosis. Neuropsychopharmacology 41: 419–430.

19. Knott V, Impey D, Choueiry J, Smith D, de la Salle S, Saghir S, et al. (2015): An acute dose, randomized trial of the effects of CDP-Choline on Mismatch Negativity (MMN) in healthy volunteers stratified by deviance detection level. Neuropsychiatr Electrophysiol 1. 10.1186/s40810-014-0002-4

20. Choueiry J, Blais CM, Shah D, Smith D, Fisher D, Labelle A, Knott V (2023): An α7 nAChR approach for the baseline-dependent modulation of deviance detection in schizophrenia: A pilot study assessing the combined effect of CDP-choline and galantamine. Journal of Psychopharmacology 37: 381–395.

21. Levinstein MR, Budinich RC, Bonaventura J, Schatzberg AF, Zarate CA, Michaelides M (2025): Redefining Ketamine Pharmacology for Antidepressant Action: Synergistic NMDA and Opioid Receptor Interactions? Am J Psychiatry 182: 247–258.

22. Gilbert JR, Galiano CS, Nugent AC, Zarate CA (2021): Ketamine and Attentional Bias Toward Emotional Faces: Dynamic Causal Modeling of Magnetoencephalographic Connectivity in Treatment-Resistant Depression. Front Psychiatry 12: 673159.

23. Knott V, Choueiry J, Dort H, Smith D, Impey D, De La Salle S, Philippe T (2014): Baseline-dependent modulating effects of nicotine on voluntary and involuntary attention measured with brain event-related P3 potentials. Pharmacol Biochem Behav 122: 107–117.

24. Martin J, Nezhad FG, Rueda A, Lee GH, Charlton CE, Soltanzadeh M, et al. (2024): Predicting treatment response to ketamine in treatment-resistant depression using auditory mismatch negativity: Study protocol. PLoS One 19. 10.1371/JOURNAL.PONE.0308413

